# Mathematical model for the mitigation of the economic effects of the Covid-19 in the Democratic Republic of the Congo

**DOI:** 10.1101/2020.12.14.20248182

**Authors:** Zirhumanana Balike Dieudonné

## Abstract

A Mathematical model for the spread of Covid-19 in Democratic Republic of Congo taking into account the vulnerability of congolese economy is proposed. The reproduction number for the Covid-19 is calculated and numerical simulations are performed using Python software. A clear advice for policymakers is deduced from the forecasting of the model.

## Introduction

In December 2019 a novel Coronavirus appeared in Wuhan, China [9]. The international committee for taxonomy of virus has attributed the name SRAS-Cov-2 to that disease [10], [4]. On January 30, 2020; the World Health Organization (WHO) declared it to be an epidemic of international concern [19]. Four months later, the virus was spread out worldwide, only less than 10 countries were not yet touched by the disease. This is why WHO declared it to be a pandemic since March 11, 2020 [19]. Countries affected by this outbreak have envisaged several measures to mitigate its negatives effects on their health care systems. Those measures include national lockdown and cancellation of travels to and from outside their borders [14]. Many low-income countries, including African countries, have also adopted the above measures without taking into account of the vulnerability of their economies which rely mainly on informal system [23].

On 10 March 2020, the first infected individual has been detected in Democratic Republic of Congo (DRC, [20]). On March 18, 2020; the DRC President has announced draconian measures to mitigate the circulation of the pandemic. These measures include cancellation of flights out and from Kinshasa (the capital city) to other provinces, cessation of schools and churches activities, suspension of barrooms and restaurants activities, prohibition of gatherings of more than twenty persons, etc. These measures have come in force since March 19, 2020 [11].

However, these measures have drastically impacted the national economy. A study in [21] shows that hunger could be deadlier than Covid-19 in low-income countries. This is exactly the case for the DRC. Individuals whose daily income rely on the informal system are not able to strictly respect them. While they were set to control the epidemic, this economic aspect makes them unappropriated to Congolese context. In the proposed model, there is no need of shutting down all the country, only suspicious and infected people are quarantined. The meaning of ‘suspicious’ will be precised in the following section.

Since the apparition of this virus, several models have been proposed [1], [2], [3], [5], [6], [7], [8] and [13]. At my knowledge, there is no model which has been performed to help understanding the spreading of this pandemic in DRC and taking into account the economic impact of the virus on the populations. To fill in this gap and help addressing this challenge, an SEIR model (see [18], [15] and [17] for details of this model) with additional compartments is proposed in this paper. In this model, full lockdown or national quarantine is not envisaged as it may jeopardize the fragile economic system of the country. The following compartments have been added to the traditional SEIR model : *Quarantined* and *Hospitalized*.

## 1. Description of the model

The population is divided into the following subgroups:

- **Susceptibles (S):** This compartment comprises individuals who are not yet infected and not immunized against the disease. They are recruited at rate *θ* and transferred into the Exposed group at the rate *β*. In DRC, only one laboratory can declare positive individuals amid a Covid-19 testing. It is the National Institute of Biomedical Research (NIBR) located in Kinshasa, the capital city. In many cases, after testing, individuals have to wait for many days before they can get the result as the NIBR is far away from many provinces (for example, Kinshasa is 2000 km from Bukavu) and the transportation system is bankrupted. To prevent potential infected individuals to spread the disease, authorities should quarantine all suspicious individuals from susceptible population at rate *E*. Suspicious individuals are those who have been in contact with an infected (confirmed by testing) person or who have recently sojourned in a high risky area. Individuals quarantined are therefore not necessarily infected but they present some like Covid-19 symptoms or they have been (or are suspected to have been) in contact with a suspicious or confirmed case.
- **Exposed (E):** This group contains individuals who are infected but not yet infectious. They move into the Infectious (I) group at rate *α*.
- **Infectious (I)**: In this group we have individuals who are infectious, i.e those who are able to spread the disease. In general such individuals are at the onset of the illness. Thus, they will be transferred into the hospital (H) at rate *ν* and are removed at rate *η*. This removal is due either to death or recovery without being transferred at the hospital. This will particularly happen for young people who are resilient to the disease and some unbelievers who never accept that the virus is deadly and are currently propagating into the population.
- **Hospitalized (H)**. Two categories of individuals are in this group: those who come from infectious group (I) and those who come from quarantine (Q). They are respectively recruited at rates *ν* and *σ*. Hospitalized individuals are likewise removed (recovery or death) at rate *µ* amid the virus infection.
- **Quarantined (Q)**: Individuals who are deemed suspicious are placed in quarantine from Susceptible group (S) at rate *E* as mentioned above. Then, individuals are transferred either in hospital at rate *σ* or returned into Susceptible group at rate *γ* if they are declared negative after testing.
- **Removed (R)**: Removed subjects are either recovered or deceased due to Covid-19.

In addition, in all compartments, individuals are removed at the same rate *λ* due to natural death. The Fig 1 depicts the structure of this model.

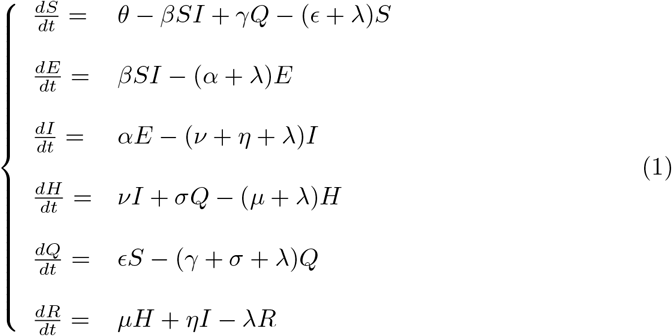

**Fig 1.**
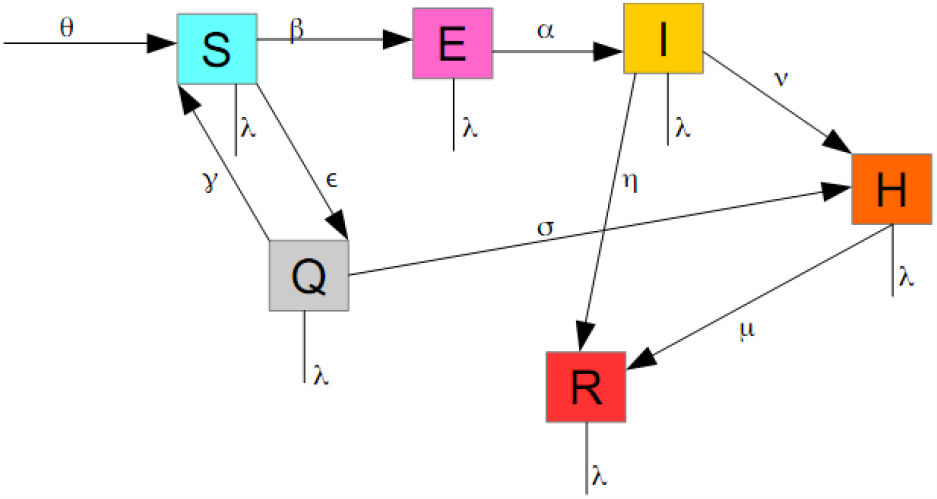
Structure of the proposed model.

In this section, an analysis of the model (system 1) will be performed.

It is clear that this system under study satisfies the Lipschitz (see [12]) and theorem 1 (see [22], page 343) conditions. This guarantees the existence and the uniqueness of the solution.

### 1.1 Feasible solution and positivity of the solution

The model is performed for understanding the spread of the SRAS-Cov-2 into a human population. Thus, the parameters are expected to be positive.

Let Ω be the set of all feasible solution to the system 1. Each solution to a single equation lie in ℝ_+_.

We hence have

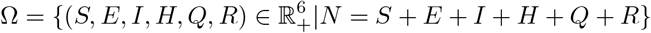

where *N* is the total size of the population under study.

In this region, I want

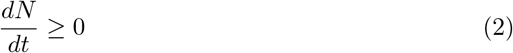

#### Proposition 1

The set Ω is invariant and attract all solutions in 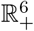.

**Proof**. From the assumption that *N* = *S* + *E* + *I* + *H* + *Q* + *R*, I deduce that

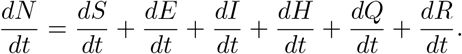

Substituting each derivative by the corresponding value from system 1 and simplifying, I get :

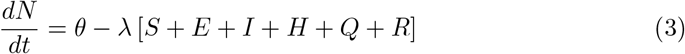

This equation can be rewritten in the following way:

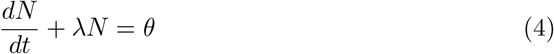

Equation 4 can be solved using an integrating factor.

Multiplying both sides of that equation by e ^*∫λdt*^, we get:

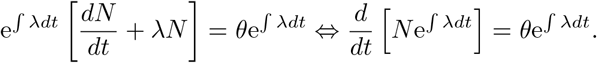

We finally get

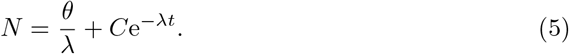

where C is an arbitrary constant. For *t* = 0, we have 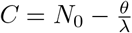 which we substitute in equation 5 to find

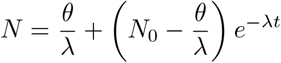

Thus, we have 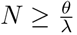 as *t →* +*∞*, provided equation 2.

This establishes that Ω is positively invariant and attracts all solutions in 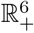.

In addition, it can be proven that the solution of system 1 has a positive solution provided the initial data set (*S*_0_, *E*_0_, *I*_0_, *H*_0_, *Q*_0_, *R*_0_) ≥0 *∈* Ω.

For example, we have

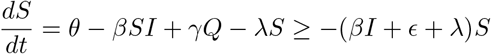

That is

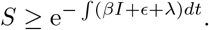

The right side of this inequality is always positive, that is the function S is also positive and consequently its initial value.

A similar computation can show that

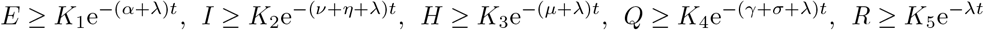

where *K*_*i*_, (*i* ∈ {1, …, 5}) are arbitrary constants.

### 1.2 The disease free equilibrium (DFE)

From the equation 1, we set *f*_*i*_ = 0, *i* = 1, …, 6 to get the new algebraic system below.

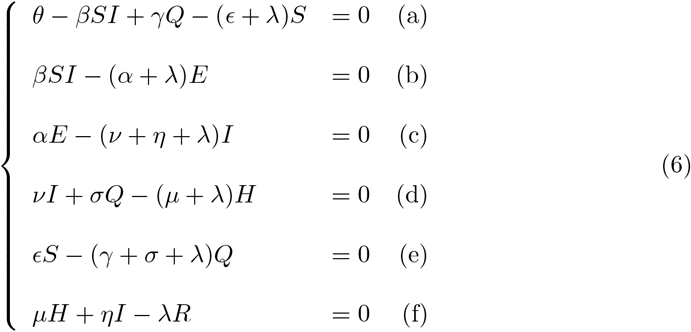

From equations (a) to (f), one successively get

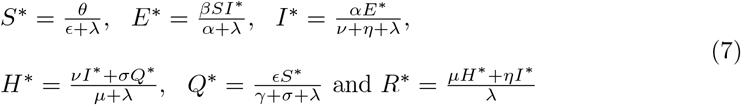

If *E* = 0 (that is *β* = 0) and *E* = 0, then *I* = *H* = *Q* = *R* = 0 and accordingly, the DFE is 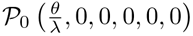. In this case, there is no disease as no infectious individuals are within the population.

If *ϵ* ≠ 0, the situation is such that suspicious are quarantined but they are neither infectious nor confirmed infected. This is consistent with the logic of the model: individuals with like Covid-19 symptoms are quarantined waiting for the testing results from the NIBR. The corresponding DFE is 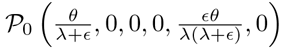, 0 as *λ* never vanishes.

### 1.3 The basic reproduction number

The infection components in this model are E, I, H and Q. In accordance with the notations in [16], [13] and [2], the new infection matrix *F* and the transition matrix *V* are given by

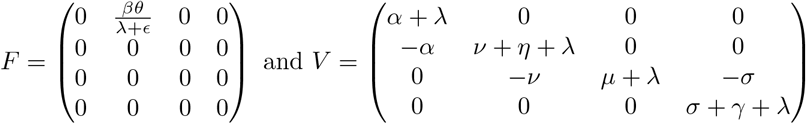

The basic reproduction number of model 1 is then defined as the spectral radius of the next generation matrix *FV* ^−1^, i.e

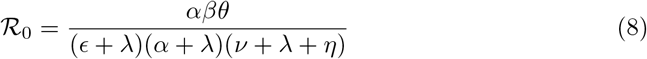

If the inequality ℛ_0_ 1 is satisfied, then no epidemic outbreak is possible, otherwise an epidemic occurs.

If *ϵ* = 0, then the fifth equation of the model 1 is removed from the infection components (that is, the model comprises only three infection components: E, I and H).

Summing equations (b) and (c) of the system 6 we get the following:

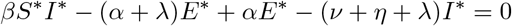

After substituting *I*^*∗*^ from equation 7 and simplifying, we get:

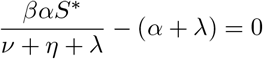

as *E*^*∗*^≠ 0.

This finally yields

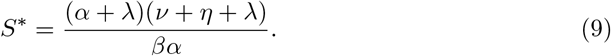

Provided 2 and its result, I have

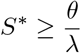

that is

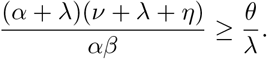

Hence, I get

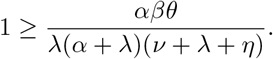

I set

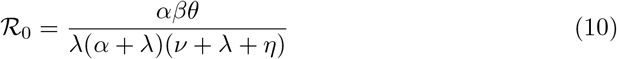

the new reproduction number.

This is exactly the spectral radius of the new next generation matrix obtained after ignoring the fifth equation of the system 1 in the infection compartments.

Thus, *ϵ* stands for a control on the spread of the disease within the susceptible population.

### 1.4 Local stability

I examine the local stability of the DFE by the mean of the jacobian matrix of the functions *f*_*i*_, *i* = 1, …, 6 (where *f*_*i*_ are the functions in the right side of the system 1) at that point. I therefore have the jacobian below

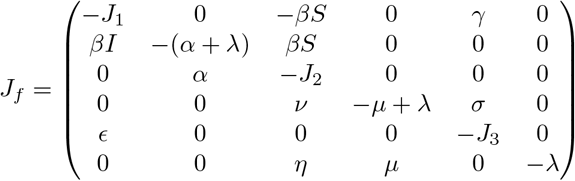

where *J*_1_ = *βI*^*∗*^ + *λ* + *E, J*_2_ = *ν* + *η* + *λ, J*_3_ = *σ* + *γ* + *λ*.

This jacobian evaluated at the DFE yields

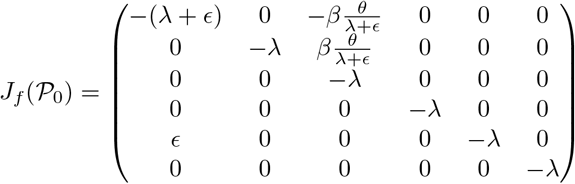

has all its eigenvalues negative whether *E* is null or not.

Thereby, the DFE is locally asymptotically stable.

## 2. Results and discussion

This section is devoted to numerical simulations and discussion of the results. One hindrance here is the lack of data related to the Covid-19 in DRC. I fitted data to a curve estimation using Python. The Fig 2 depicts how my model fits the data collected for the 100 first days of Covid-19 crisis in DRC 25.

**Fig 2.**
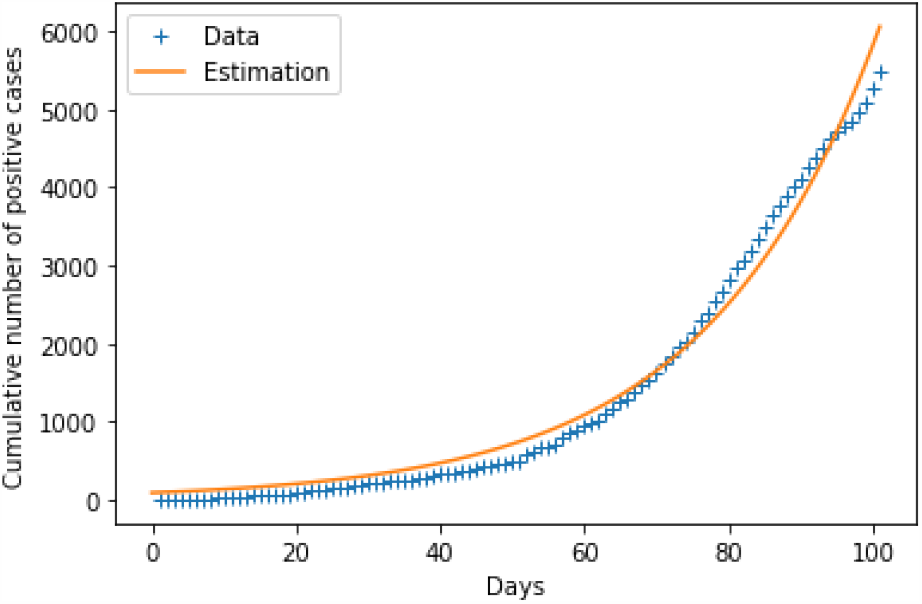
Fitting data to the model for parameters estimation

Due to lack of available data in DRC, some parameters have been reasonably assumed (without either overlooking or exaggerating them). Provided some parameters after fitting the model with data, some other parameters have been calculated and others were borrowed to [2]. To determine the value of the calculated parameters, I proceed in the following way: At the beginning of the epidemic, *E* = *I* and second equation of system 1 becomes

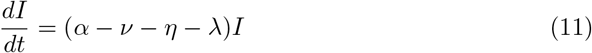

whose solution is

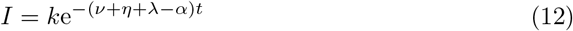

where *k* is an arbitrary constant. Using the value got from data fitting, I get

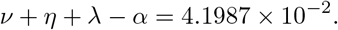

Since the value of *λ* is known from the data on life expectancy in DRC (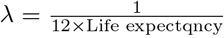, see [15] and [24] for details) and provided there some others parameters borrowed from [2], I get the values in Table 1.

**Table 1.** Parameters used in the model

| Parameters | Description | Values | Sources |
| --- | --- | --- | --- |
| $\theta$ | Population influx rate | $271.4 \text{ day}^{-1}$ | [2] |
| $\beta$ | Transmission rate | $5445 \times 10^{-6} \text{ pers}^{-1} \text{ day}^{-1}$ | Calculated |
| $\frac{1}{\alpha}$ | Incubation period | 14 days | [2] |
| $\lambda$ | Natural death | $0.001378 \text{ pers day}^{-1}$ | Calculated |
| $\nu$ | Hospitalization rate | $0.0097 \text{ person day}^{-1}$ | Assumed |
| $\eta$ | Removal (recovery and death) rate | $0.01428 \text{ pers day}^{-1}$ | Data |
| $\sigma$ | Transfer rate from Q to H | $\frac{1}{14} \text{ day}^{-1}$ | Ref |
| $\epsilon$ | Quarantine rate | Variable $\text{pers day}^{-1}$ | Assumed |
| $\gamma$ | Transfer rate from Q to S | $\frac{1}{14} \text{ day}^{-1}$ | Assumed |
| $\mu$ | Removal rate from hospital | $\frac{1}{28} \text{ day}^{-1}$ | Assumed |

Using the values of parameters like provided in Table 1, a simulation of the model has been performed in Python. The model is designed such that Susceptible stratum collapses as early as possible. This makes the Exposed and infectious groups to explode quickly.

The impact of *E* can be evaluated on Fig 3 and Fig 4. When the number of individuals in group Q is high, the number of positive cases drops but the number of individuals hospitalized increases. This is a proof that organized quarantine allows a rapid detection of positive cases and enables the timely management of patients who need to be hospitalized. The fact that the infectious curve reaches its peak without ever affecting the maximum of the population under study is proof that many individuals are freely circulating as long as they have not been deemed suspicious for quarantine. This allows the economy to run normally in the midst of a virus crisis.

**Fig 3.**
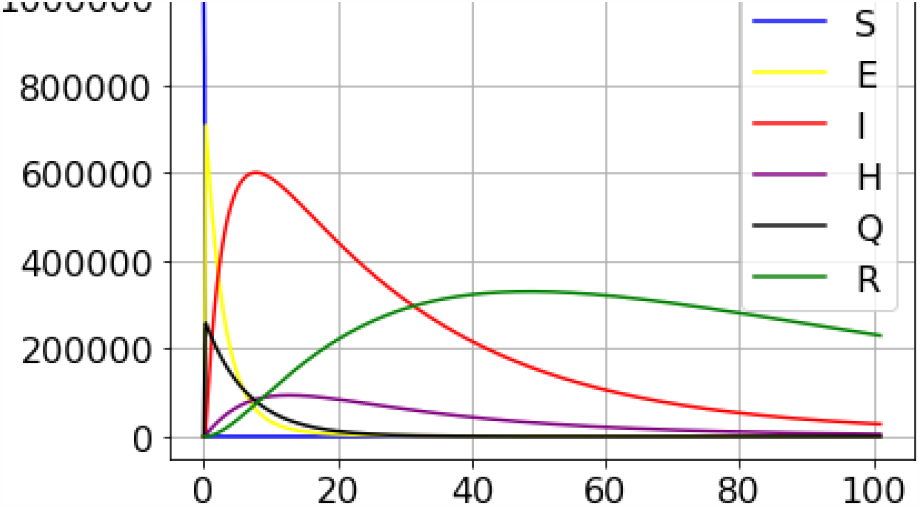
A simulation result for the outbreak using data from the 100 first days data from DRC (*ϵ* = 1.5).

**Fig 4.**
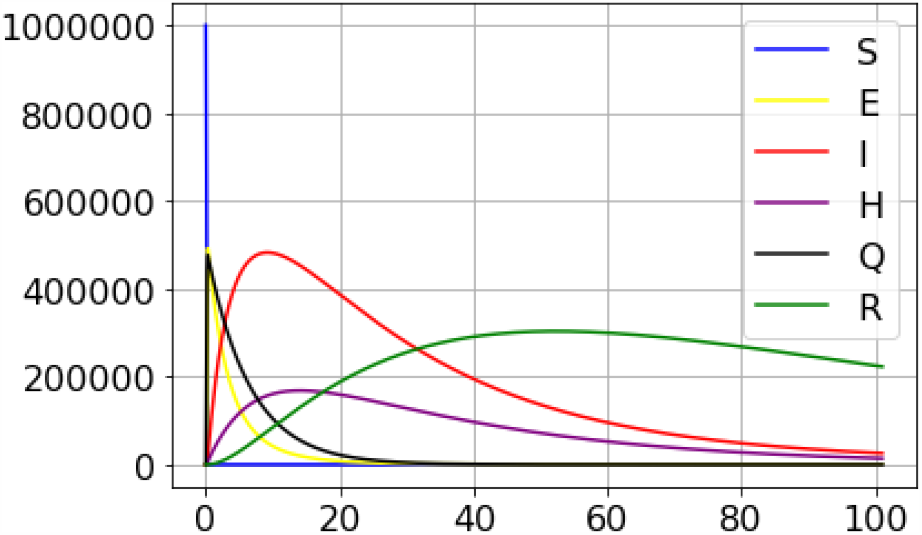
A simulation result for the outbreak using data from the 100 first days data from DRC(*ϵ* = 3.1).

## 3 Conclusion

The mathematical model proposed in this paper has been designed taking into account the vulnerability of the economy of the Democratic Republic of the Congo and other low-income countries. Since the country does not have enough means to control the epidemic from Kinshasa as is the case to date, I propose that the management of this crisis be decentralized. Thus, each provincial governor will be able to set up a team responsible for tracking down suspect people and quarantine them while waiting for the results of the NIBR tests.

## Data Availability

Public data, access guranteed

http://www.covid19drc.com

## Notes

### Competing Interest Statement

The authors have declared no competing interest.

### Funding Statement

Not applicable: no funding has been provided.

